# Fatty acid profiles, cholesterol composition, and nutritional quality indices of 37 commonly consumed local foods in Kuwait in relation to cardiovascular health

**DOI:** 10.1101/2020.11.18.20233999

**Authors:** Hanan A. Al-Amiri, Nisar Ahmed, Tahani Al-Sharrah

**Affiliations:** Food and Nutrition Program, Environment and Life Sciences Research Center, Kuwait Institute for Scientific Research, Shuwaikh, Kuwait; Environment Pollution and Climate Program, Environment and Life Sciences Research Center, Kuwait Institute for Scientific Research, Shuwaikh, Kuwait

## Abstract

Dietary fatty acids and cholesterol content are proved, by many research studies, to be associated with various health conditions, including cardiovascular health. Knowledge of the composition of these nutrients in food is essential for proper planning of health programs. The present study aimed at assessing the fatty acid profile, fatty acid nutritional quality, and cholesterol composition of 37 foods commonly consumed in the State of Kuwait and the potential impact of these foods on cardiovascular risk. Fatty acid profile was determined by gas chromatography-flame ionization detector into four types: saturated, monounsaturated, polyunsaturated, and trans fatty acids. Nutritional quality was calculated using the atherogenic index, thrombogenic index, hypocholesterolemic/hypercholesterolemic fatty acid ratio, polyunsaturated fatty acid/ saturated fatty acid ratio, and n-3/n-6 fatty acids ratio. Determination of cholesterol was performed by gas chromatography. Saturated fatty acid levels ranged from 0.01–21.83, monounsaturated fatty acids 0.01–25.51, and 0.013–22.87 g/100 g edible portion of food. The predominant fatty acids identified in all studied foods were C18:2c (n-3), C16:0, and C18:1c, with values 0.45–56.52, 10.12–44.90, and 16.99–42.56% of total fatty acids, respectively. The trans fatty acid content was low in all foods. Cholesterol levels varied between traces (<0.05) and 454.79 mg/100 g edible portions of food. Results show that seafood, rice-based, seed-based, and vegetable-based foods had better nutritional quality in terms of the fatty acids content, as indicated by the polyunsaturated fatty acid/saturated fatty acid and n-3/n-6 ratios, low thrombogenicity indices, and high hypocholesterolemic/hypercholesterolemic fatty acid ratios. Cholesterol and fatty acid data obtained in the present study will be of special interest for many studies, including nutrition-related health research, and will help policymakers in proper strategies for health programs.

## Introduction

Dietary intake of fatty acids and cholesterol play an important role in the etiology of cardiovascular disease (CVD) [1, 2], a leading cause of death in most countries [3]. According to the World Health Organization (WHO), CVD accounts for 17.9 million deaths annually [3].

Several experimental and epidemiological studies have proven the strong association between specific types of dietary fatty acids and blood cholesterol levels and therefore, the risk of CVD [2, 4-8]. The degree of CVD risk may vary according to the type and level of dietary fatty acids. Dietary saturated fatty acid (SFA), trans fatty acid (TFA), and/or improper ratio of n-3/n-6 fatty acids are common dietary causes of CVD [6, 9]. Dietary SFAs elevate total and low-density lipoprotein (LDL) blood cholesterol, whereas TFAs elevate total blood cholesterol and LDL cholesterol, and lower blood high-density lipoprotein (HDL) cholesterol [9, 10]. However, dietary polyunsaturated fatty acid (PUFA) and monounsaturated fatty acid (MUFA) generally lower both total and LDL cholesterol levels, thus may cause reduction in the incidence of CVD [11, 12], and possibly the prevention of some cancers, asthma, diabetes and hypertension [13]. Additionally, a strong relationship has been revealed between cellular cholesterol levels and Alzheimer’s disease [14]. Nutritional strategies to achieve a better balance of fatty acids in the diet by decreasing the intake of SFA and TFA as well as cholesterol, appear to be significant and effective to reduce the potential risk of these diseases. Thus, knowledge of the fatty acid and cholesterol composition of foods is essential before any nutritional strategy can be implemented.

This article aims to provide detailed data on the fatty acid profiles and nutritional quality, as well as cholesterol contents of 37 local foods commonly consumed in the State of Kuwait. This data will contribute valuable information to fatty acid and cholesterol composition tables. The enrichment of food composition tables with reliable analytical data on local foods is significant for obtaining convincing results in future nutritional-health related research and in the development and implementation of nutritional policies aimed at improving health.

## Materials and methods

### Food sampling

Top thirty seven most commonly consumed local foods among Kuwaitis were selected from “Kuwait Total Diet Study” data. The selected foods included 21 foods commonly cooked and prepared at home, and 16 ready-made foods often purchased by locals rather than cooked at home. Each ready-made food was purchased from three different restaurants/confectioneries based on its popularity among Kuwaitis. The most common brands of cream cheese spread (four brands), triangle processed cheese (three brands), and white cheese low fat (one brand “Al-Wafra”) were purchased from retailers.

As for cooked foods, data on the ingredients, their quantities, and cooking procedures were collected based on a field survey of 250 Kuwaiti households. The food recipes were standardized by identifying the foremost ingredients of each recipe and their weight ratio from the total, ensuring that the coefficient of variation (CV) did not exceed 20%. Details on the major ingredients of the 37 foods are given in S1 Table.

The cooked foods were prepared at the Ministry of Health (MOH) kitchen based on the standardized recipes under the supervision of experienced Kuwaiti cooks. All ingredients including water were weighed, and three identical preparations of each food were cooked separately. Foods were cleared of inedible parts, if any, including bones and skin. Each cooked and ready-made food was then homogenized thoroughly in a blender and sampled for moisture analysis. The remaining food sample was dried in a freeze drier (VirTis, Unitop 800L, USA), ground, transferred into airtight bottle containers, labeled, and stored in a deep freezer at -18°C until analysis.

### Fatty acid analysis

Total fat content in g/100 g edible portion (EP) of food was determined gravimetrically following Soxhlet extraction (Tecator Soxtec system, HT 1043 extraction unit, Herndon, VA, USA) according to the method given in AOAC [15]. Fatty acids were extracted from each sample in triplicate according to Folch, Lees, and Sloane Stanley [16] with a modification by Segura and Lopez-Bote [17]. Briefly, 0.5 g of freeze dried food sample was homogenized with 20 ml chloroform:methanol (2:1 v/v) solution. The samples were ultrasonicated at room temperature for 15 min. After allowing to settle for a few minutes, the homogenate was filtered. The filtrate was collected in a tube and NaCl solution was added. The tube centrifuged for 20 min at approximately 3000 x rmp. The upper layer of methanol:water was removed, and the lower layer was filtered through anhydrous sodium sulphate and collected in a volumetric flask. The solvent was evaporated under nitrogen, the residual fat was esterified, and methyl esters of fatty acids (FAME) were prepared as described by the American Oil Chemists’ Society (AOCS) Official Method Ce 2-66 [18]. A 0.5 μL esterified sample was injected into a gas chromatograph (GC) (Agilent 6890 Series, Agilent Technologies, Palo Alto, CA, USA) coupled with a flame ionization detector (FID). A cyano-polysiloxane (CP-Sil 88 for FAME, part number CP7489) capillary column (100 m × 0.25 mm, 0.20 µm; Agilent Technologies, Palo Alto, CA, USA) was used for the FAMEs separation under the following instrumental conditions: injector and FID detector temperatures were 260 and 270°C, respectively, with injector split ratio of 1:50, and carrier gas helium with a constant flow rate of 1.0 mL/min. The initial oven temperature was 80 °C, at which it was held for 5 min, followed by an increase to 220 °C at a rate of 4 °C/min then was held for 5 min, and finally the temperature was increased to 240 °C at a rate of 1 °C/min and was held for 10 min. A 37-component FAMEs standard mixture was used to identify the fatty acids, the mixture contained FAMEs ranging from C4:0 to C22:6. The FAMEs were identified by comparing the retention times of standard peaks with the retention times of sample peaks. The quantification of the fatty acids was performed by the conversion of the peak areas in percentage of the extract using a GC Workstation software. The fatty acid content was reported as percentage of the total fatty acids. In order to compile data in food composition tables, fatty acid data need to be transformed to per 100 g EP on fresh weight basis, to create internationally interchangeable food composition data. The fatty acid composition in g/100 g EP was calculated from the corresponding fatty acid (percentage of total fatty acids), following the method described by Paul and Southgate [19].

### Cholesterol analysis

Cholesterol was analyzed in triplicate via GC (Agilent 6890 Series, Agilent Technologies, Palo Alto, CA, USA) according to a method described by Naeemi et al. [20]. Each sample was spiked with the internal standard 5-α-cholestane after saponification with a saturated methanolic KOH solution; hexane was then added, centrifuged at 2000 rpm for 2 min, and finally 0.5 μL of the upper layer was injected into the GC for cholesterol analysis.

The GC conditions used in this study were as follows: injector and detector temperatures were 300°C, with injector split ratio of 1:25, and carrier gas helium with flow rate of 1.0 mL/min. The initial oven temperature was 250 °C, the holding time was 2 min, and the temperature rise rate was 3 °C/min.

### Fatty acid quality

Fatty acid quality was determined using the atherogenic index (AI) and thrombogenic index (TI) according to Ulbricht & Southgate [21]. The hypocholesterolemic to hypercholesterolemic (H/H) fatty acid ratio index which considers the specific effects of fatty acids on cholesterol metabolism, was determined as described by Testi et al. [22]. The indices were calculated using the following equations:

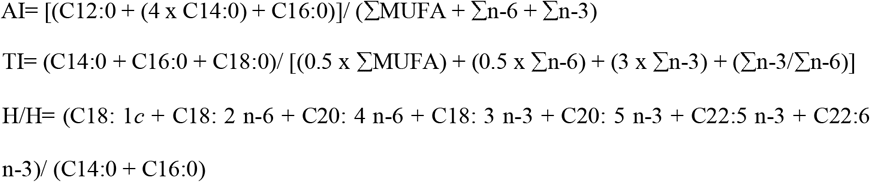

where:

∑MUFA = sum of monounsaturated fatty acids,

∑n-6 = sum of n-6 fatty acids, and

∑n-3 = sum of n-3 fatty acids.

Further indices were calculated as PUFA to SFA (P/S) ratio, and n-3/n-6 fatty acids ratio.

### Statistical analysis

Each food sample was analyzed in triplicate for total lipid, fatty acid, and cholesterol composition. Fatty acid data were reported as the mean value on the wet weight of the edible portion of food in g/100 g, and as percentage of the total fatty acids. Cholesterol data were reported as the mean ± standard deviation (SD) in mg/100 g EP of food. The minimum and maximum values for each fatty acid and cholesterol content were interpreted as a measure of range.

## Results and discussion

### Fatty acid profile

Investigated foods in the current study were classified into nine categories: seafood (n=3), soup (n=3), cheese (n=3), rice-based (n=6), desserts (n=11), seed-based (n=2), meat-based (n=4), sandwiches (n=2), and vegetable-based (n=3) (Table 1). The classification of the foods is based on major ingredients in each recipe, as well as the common classification (i.e. cheese, soup, desserts, and sandwiches). All cheese, desserts (except *Khabeesa* “cooked semolina with oil and sugar” and *Aseeda* “cooked wheat flour with sugar”), sandwiches, one meat-based food (*Arayes* “pita bread stuffed with minced lamb”), and one seed-based food (*Harda* “soft sesame butter, tahini”) were ready-made and purchased from the most popular restaurants and bakeries/confectioneries. The detected fatty acids were classified into four types: SFA, MUFA, PUFA, and TFA. The fatty acid compositions were expressed both as g/100 g EP and as % of the total fatty acids.

**Table 1.**
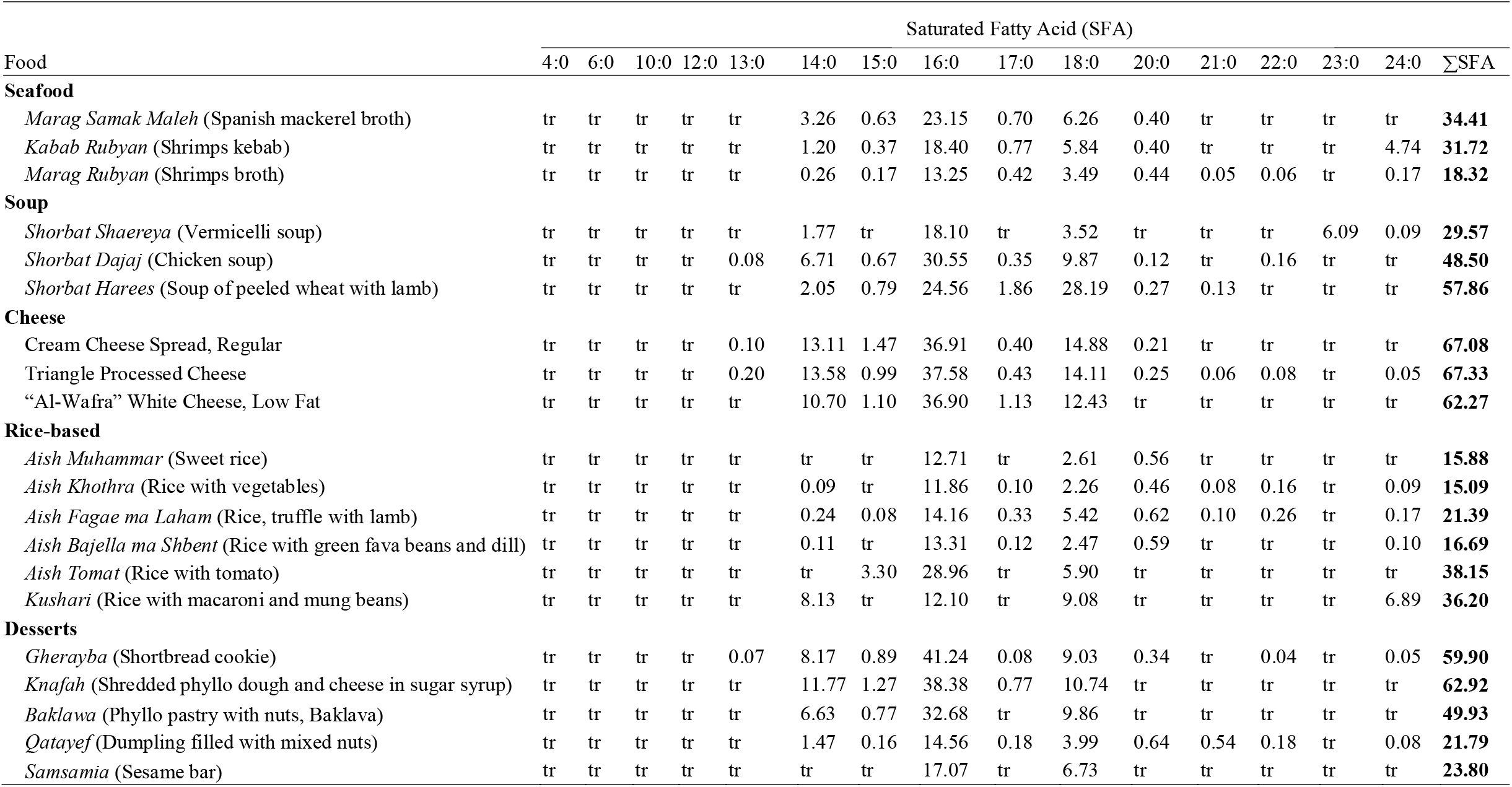
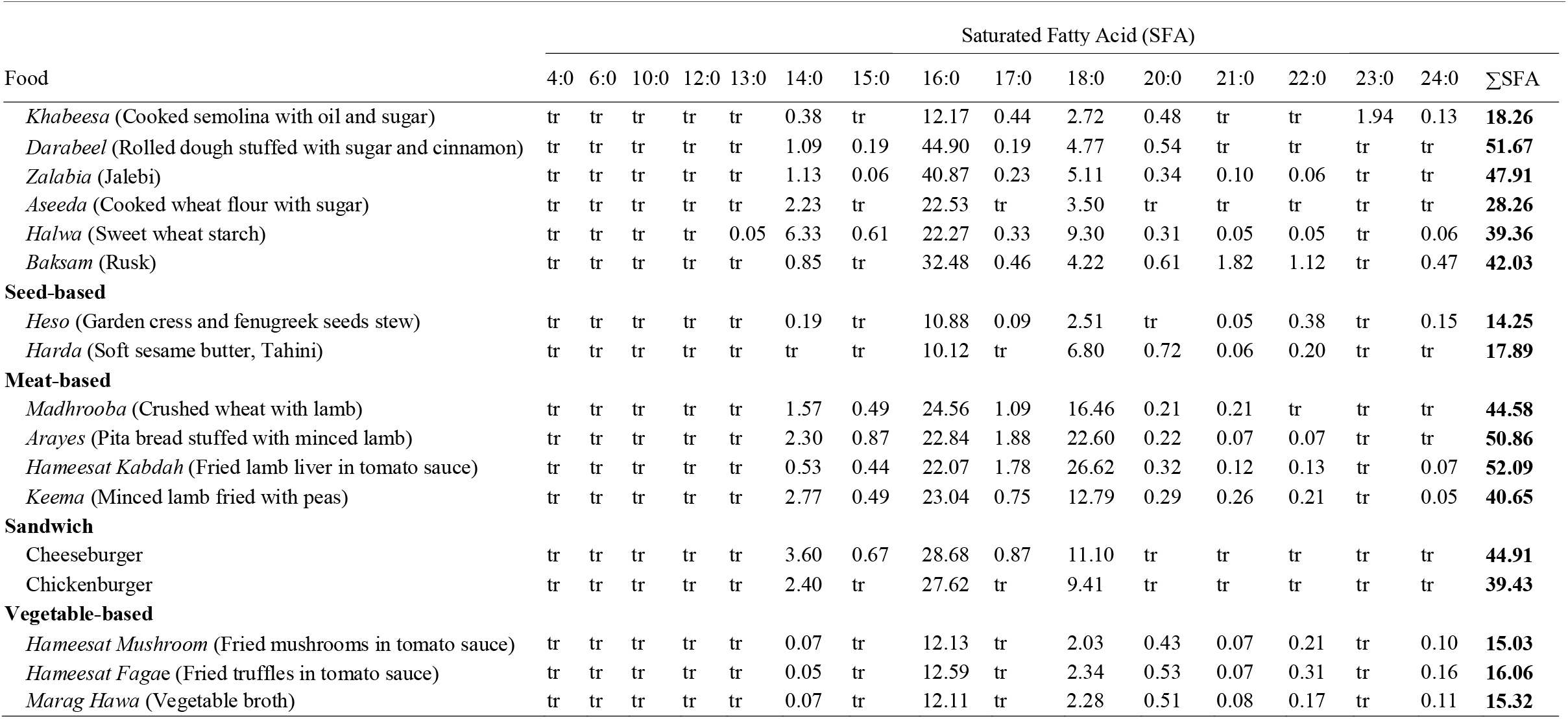
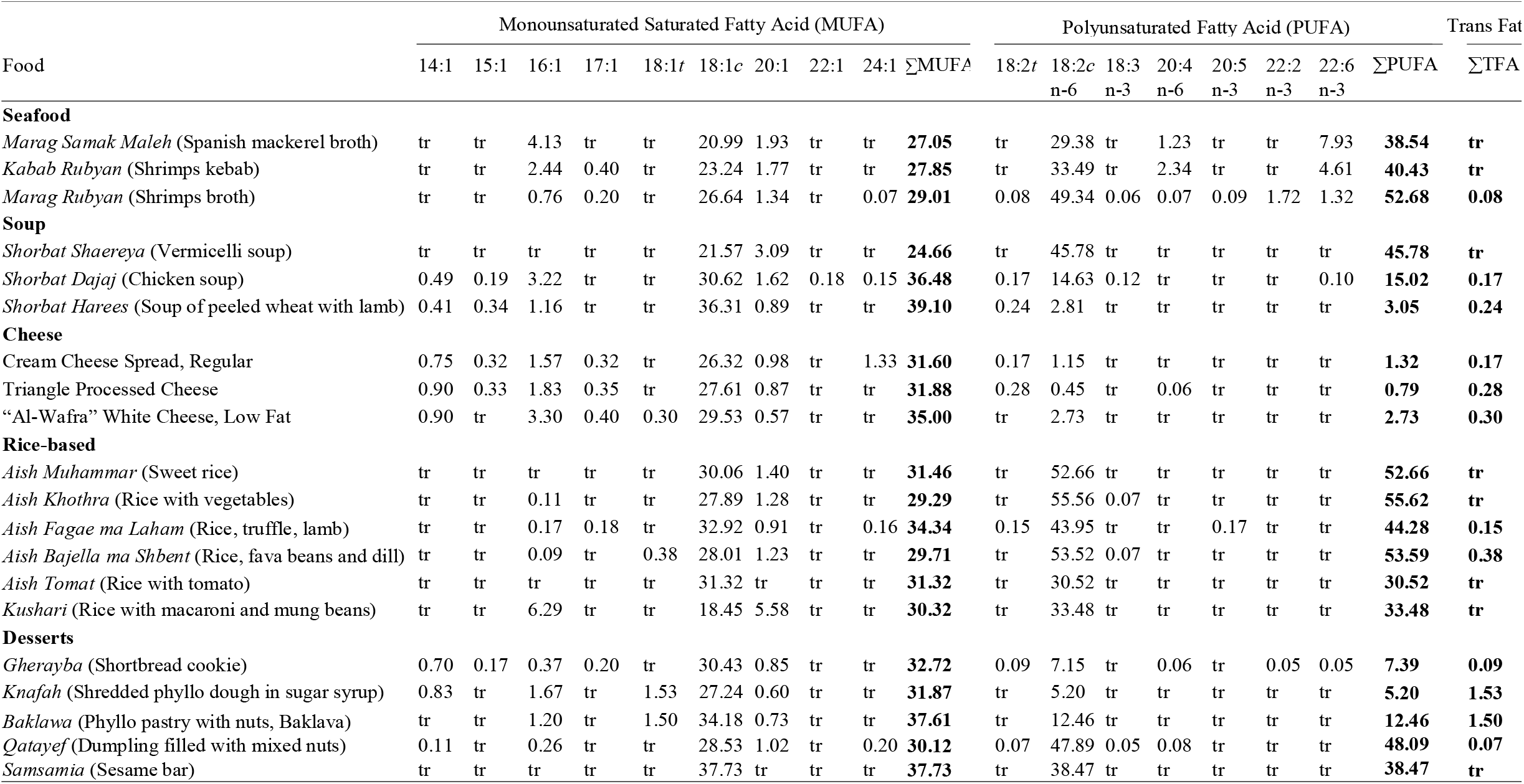
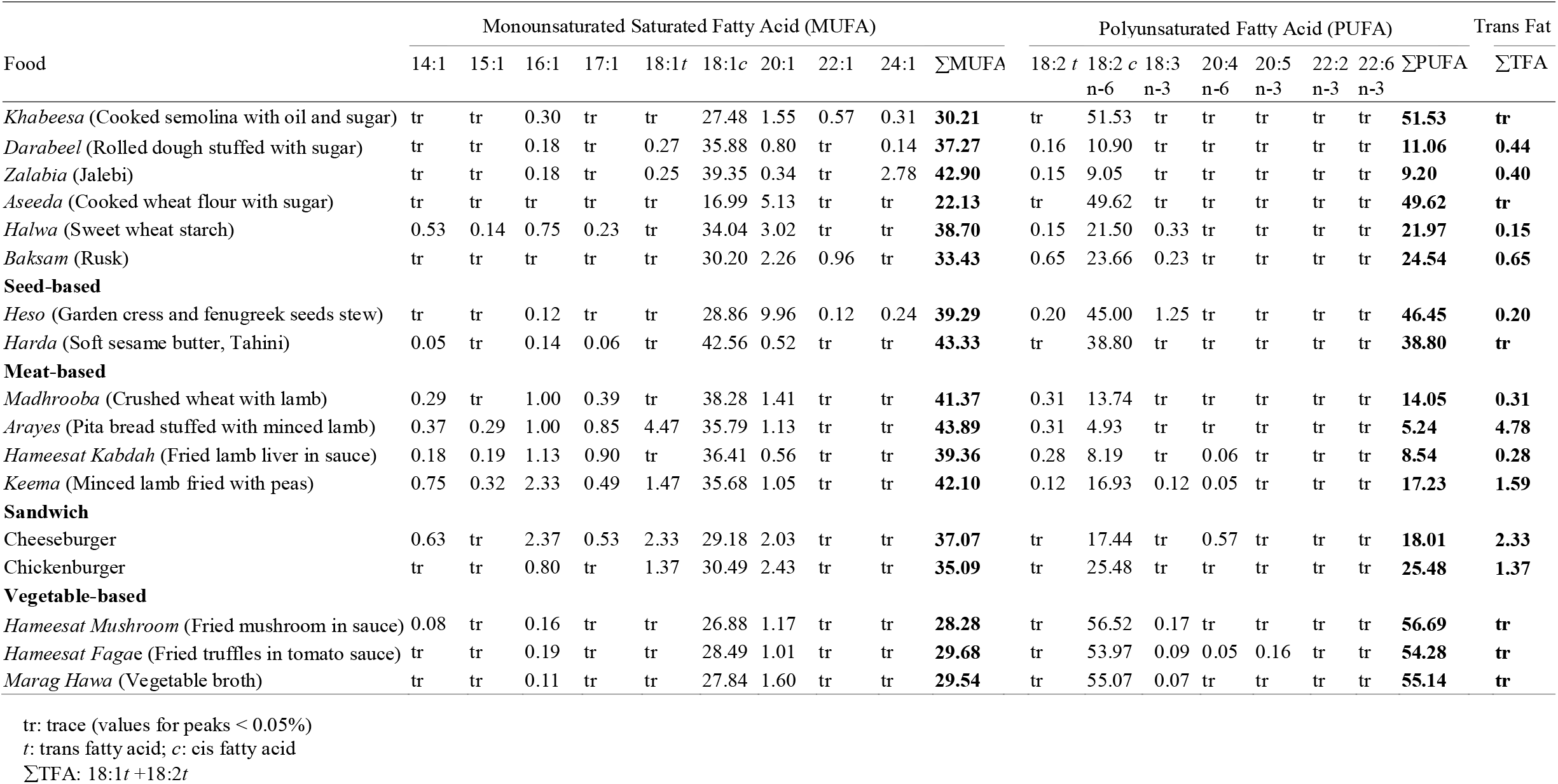
Fatty acid composition profile on the wet weight of the edible portion (percentage of the total fatty acids) for 37 commonly consumed local foods in the State of Kuwait

### Saturated fatty acids

SFA was the most dominant type of fatty acids among analyzed foods (Fig 1). The SFA content was observed with values ranged from 0.01 to 21.85 g/100 g EP (S2 Table). The highest range of SFA content (8.32–21.85 g/100 g EP) and percentage of total fatty acids (62.27–67.33%) was found in cheese. Meat-based foods and sandwiches showed high levels of SFA ranged from 40.65 to 52.09, and 39.43 to 44.91% of total fatty acids, respectively. *Knafah* (shredded phyllo dough and cheese in sugar syrup) and *Gherayba* (shortbread cookie) showed high SFA concentrations of 10.96 and 15.95 g/100 g EP, respectively, and high SFA percentage of total fatty acids of 62.92 and 59.98%, respectively (S2 Table and Table 1). The high SFA content of the dessert *Gherayba* is attributed to the use of ghee in the recipe and to the use of cheese and butter in *Knafah*.

**Fig 1.**
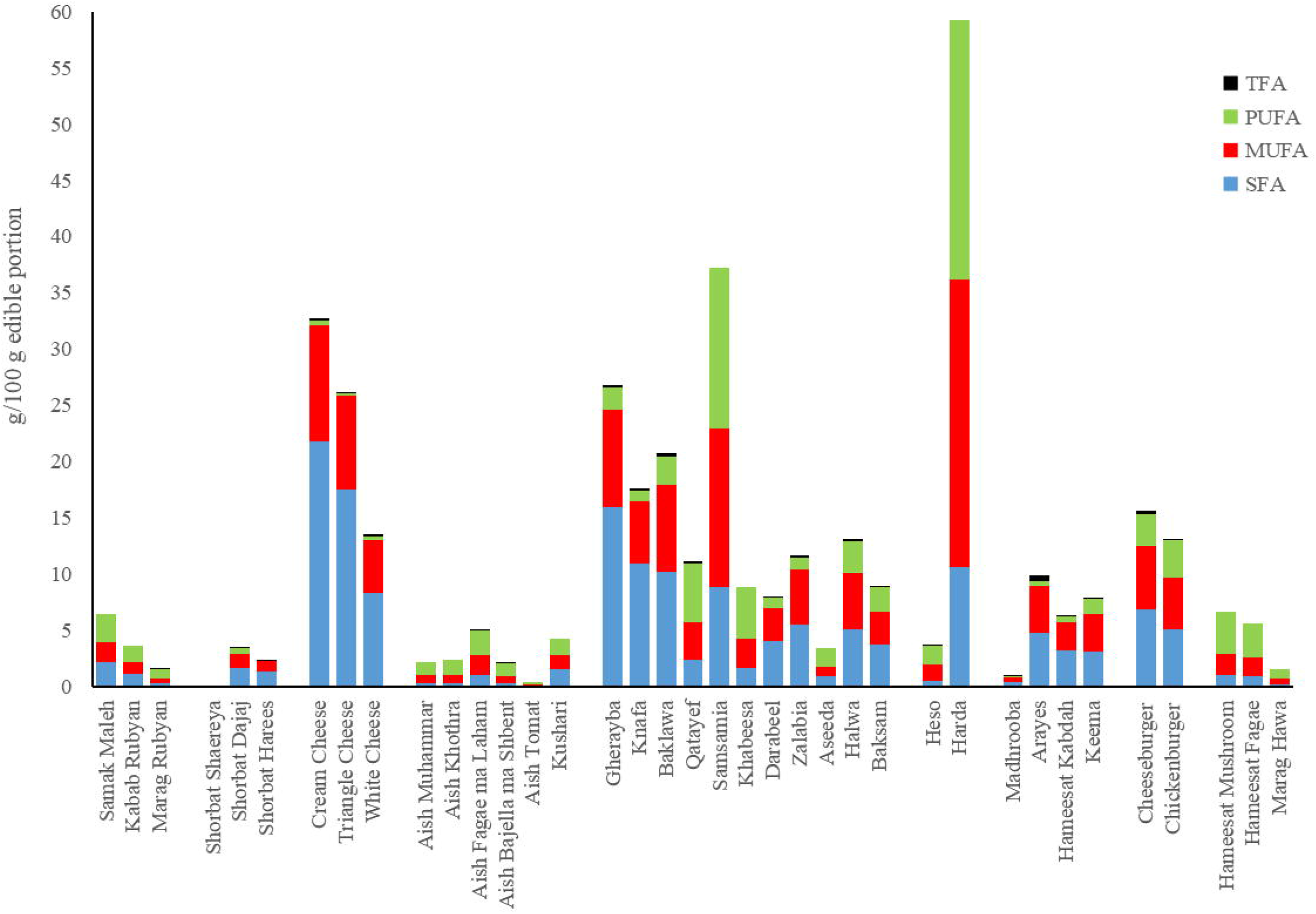
Concentrations in g/100 g edible portion of food: SFA (saturated fatty acid); MUFA (monounsaturated fatty acid); PUFA (polyunsaturated fatty acid); TFA (trans fatty acid); of 37 commonly consumed local foods in Kuwait. SFA was the most dominant type of fatty acids among foods, while TFA was found in very little amounts.

According to the joint Food and Agriculture Organization (FAO)/WHO [23] and FAO [24] reports, the SFAs with the most determinant effects on blood LDL cholesterol elevation are lauric (C12:0), myristic (C14:0), and to some extent palmitic (C16:0) acids, whereas stearic acid (C18:0) has been shown to reduce total blood cholesterol levels. Lauric acid was detected in trace amounts in all food samples (<0.05% of total fatty acids), whereas, C14:0 proportion was low with values ranging between trace amounts (<0.05%) and 8.2% of total fatty acids, except for cheese dishes (10.70–13.58%) and *Knafah* (11.77%) (Table 1). C16:0 was the most abundant SFA in 35 of 37 foods. The highest and lowest concentrations of C16:0, 12.02 and 0.01 g/100 g EP, were detected in cream cheese and *Shorbat Shaereya* (vermicelli soup), respectively (S2 Table). In contrast, the highest and lowest percentage of palmitic acid of the total fatty acids were identified in dessert *Darabeel* (rolled dough stuffed with sugar and cinnamon; 44.90%) and seed-based food *Harda* (10.12%), respectively. The C16:0 percent of the total fatty acids was predominant in cheese dishes (36.90–37.58%) and in the following desserts: *Baksam* (rusk; 32.48%), *Baklawa* (baklava; 32.68%), *Knafah* (38.38%), *Zalabia* (jalebi; 40.87%), and *Gherayba* (41.24%) (Table 1). Thus, we recommend that these foods be consumed in moderation or the butter and ghee used in desserts preparation can be substituted with unsaturated fatty acids oil. The dominance of SFA in the analyzed foods and of palmitic acid in SFA is consistent with earlier studies on similar groups of foods from Saudi Arabia [25].

### Monounsaturated fatty acids

The highest concentration of MUFA (25.66 g/100 g EP) was demonstrated by *Harda*, followed by *Samsamia* (sesame bar; 14.07 g/100 g EP) and cream cheese (10.29 g/100 g EP) (S2 Table). MUFA was the most dominant type of fatty acids in only two food samples *Keema* (minced lamb with peas) and *Harda* (42.10 and 43.33%, respectively). Oleic acid (C18:1*c*) was the most dominant MUFA across all 37 food samples. The oleic acid percentage of total fatty acids was the highest in *Harda* (42.56%), the major ingredient of this food is sesame seeds, known to be high in oleic acid [26].

### Polyunsaturated fatty acids

PUFA reduces blood LDL cholesterol, but has an undesirable effect of also reducing HDL cholesterol [11]. As presented in Table 1, PUFA was the most dominant type of fatty acids in vegetable-based foods (54.28–56.69% of total fatty acids), rice-based foods (44.28–55.62% of total fatty acids), and seafood dishes (38.54–52.68% of total fatty acids). Among PUFA, the concentration of linoleic acid (C18:2*c n-6*) was the highest in all foods, with *Harda* (22.98 g/100 g EP) and *Samsamia* (14.35 g/100 g EP) containing the highest levels which were attributed to sesame seeds constituting the major ingredient in *Harda* and *Samsamia* recipes. Linoleic acid was the dominant fatty acid in 17 of the 37 foods, which is nutritionally desirable owing to its metabolism at the tissue level, producing hormone-like prostaglandins that aid in reducing blood pressure, preventing gastric ulcers, and relieving nasal congestion and asthma, as well as diminishing platelet aggregation, thus preventing CVD [27]. Linolenic (C18:3 *n-3*), eicosapentaenoic (C20:5 *n-3*), and docosadienoic (C22:2 *n-3*) acids were detected in small quantities (<0.05 g/100 g EP) in all foods. Arachidonic (C20:4 *n-6*) and docosahexaenoic (DHA; C22:6 *n-3*) acids were detected in seafood dishes at concentrations of 0.001–0.09 and 0.02–0.51 g/100 g EP, respectively (S2 Table). The arachidonic acid and DHA percentage of total fatty acids detected in seafood dishes varied between traces–2.34 and traces–7.93%, respectively (Table 1). The availability of DHA in seafood dishes is in agreement with other studies [28, 29] pointed to the high levels of DHA in fish.

### Trans fatty acids

Trans fats are commonly found in highly processed foods, although natural trans fats produced in animals and foods made from these animals (e.g. milk, meat products) may also contain small amounts of these fats. TFA content investigated in the food samples ranged between traces and 0.45 g/100 g EP, all of which were in ready-made purchased foods and foods of animal origin. The highest TFA concentration (0.45 g/100 g EP) was in a meat-based food *Arayes*, followed by cheeseburger (0.36 g/100 g EP), and in desserts *Baklawa* (0.31 g/100 g EP) and *Knafah* (0.27 g/100 g EP) (S2 Table). The TFA percentage of total fatty acids for the same foods was the highest, with values of 4.78, 2.33, 1.50, and 1.53%, respectively (Table 1). The high TFA content in *Arayes* and cheeseburger could be attributed to the major ingredient of both of these foods being lamb and beef, respectively; whereas in *Baklawa* and *Knafah*, it could be attributed to the use of butter in *Baklawa*, and butter and cheese in *Knafah*.

In 2017, Saudi Arabia; a Middle East country with similar lifestyles, dietary patterns, eating habits, nutrition-related health concerns, and nature of foods as the State of Kuwait; implemented 2% and 5% TFA limits for fats and all other foods, respectively [30]. In light of this fact, it is interesting to note that none of the foods analyzed in the present study had TFA >5% of the total fatty acids, whereas the meat-based foods *Arayes* and cheeseburger had TFA proportion of total fatty acids exceeding 2%.

Overall, TFA has no known health benefits. There is a common understanding that high consumption of TFA has negative health effects, and has been strongly associated with a high risk of CVD [24]. TFA has worse effects on blood cholesterol levels than SFA as it not only raises LDL cholesterol but also lowers HDL cholesterol.

### Cholesterol composition

The cholesterol content of the investigated foods varied between 4.42 and 454.79 mg/100 g EP of food (Table 2). Cholesterol was not detected in 19 foods, all of which contained no animal fat, the source of dietary cholesterol, detection limit was <0.05 mg/100 g.

**Table 2.** Cholesterol composition given in means ± SD (mg/100 g edible portion of food)

| Food |  |
| --- | --- |
| **Seafood** |  |
| *Marag Samak Maleh* (Spanish mackerel broth) | 86.75 ± 4.67 |
| *Kabab Rubyan* (Shrimps kebab) | 359.17 ± 8.04 |
| *Marag Rubyan* (Shrimps broth) | 212.58 ± 6.34 |
| **Soup** |  |
| *Shorbat Shaereya* (Vermicelli soup) | ND |
| *Shorbat Dajaj* (Chicken soup) | 103.47 ± 5.56 |
| *Shorbat Harees* (Soup of peeled wheat with lamb) | 23.80 ± 0.64 |
| **Cheese** |  |
| Cream Cheese Spread, Regular | 169.16 ± 5.67 |
| Triangle Processed Cheese | 145.16 ± 6.67 |
| “Al-Wafra” White Cheese, Low Fat | 97.13 ± 4.65 |
| **Rice-based** |  |
| *Aish Muhammar* (Sweet rice) | ND |
| *Aish Khothra* (Rice with vegetables) | ND |
| *Aish Fagae ma Laham* (Rice, truffle with lamb) | 15.18 ± 2.46 |
| *Aish Bajella ma Shbent* (Rice with green fava beans and dill) | ND |
| *Aish Tomat* (Rice with tomato) | ND |
| *Kushari* (Rice with macaroni and mung beans) | ND |
| **Desserts** |  |
| *Gherayba* (Shortbread cookie) | 10.10 ± 1.49 |
| *Knafah* (Shredded phyllo dough and cheese in sugar syrup) | 56.60 ± 4.63 |
| *Baklava* (Phyllo pastry with nuts, Baklava) | 4.42 ± 0.37 |
| *Qatayef* (Dumpling filled with mixed nuts) | 15.32 ± 1.75 |
| *Samsamia* (Sesame bar) | ND |
| *Khabeesa* (Cooked semolina with oil and sugar) | ND |
| *Darabeel* (Rolled dough stuffed with sugar and cinnamon) | ND |
| *Zalabia* (Jalebi) | ND |
| *Aseeda* (Cooked wheat flour with sugar) | ND |
| *Halwa* (Sweet wheat starch) | 33.21 ± 1.25 |
| *Baksam* (Rusk) | ND |
| **Seed-based** |  |
| *Heso* (Garden cress and fenugreek seeds stew) | ND |
| *Harda* (Soft sesame butter, Tahini) | ND |
| **Meat-based** |  |
| *Madhrooba* (Crushed wheat with lamb) | 19.78 ± 0.65 |
| *Arayes* (Pita bread stuffed with minced lamb) | 67.85 ± 2.46 |
| *Hameesat Kabdah* (Fried lamb liver in tomato sauce) | 454.79 ± 31.01 |
| *Keema* (Minced lamb fried with peas) | 72.42 ± 2.97 |
| **Sandwich** |  |
| Cheeseburger | 123.42 ± 1.71 |
| Chickenburger | 68.00 ± 7.77 |
| **Vegetable-based** |  |
| *Hameesat Mushroom* (Fried mushrooms in tomato sauce) | ND |
| *Hameesat Fagae* (Fried truffles in tomato sauce) | ND |
| *Marag Hawa* (Vegetable broth) | ND |

SD: standard deviation
ND: Not detected (detection limit <0.05 mg/100 g edible portion of food)

The highest cholesterol content of 454.79 mg/100 g was found in *Hameesat Kabdah*. This food is based on lamb liver that is known to be rich in cholesterol with 386 mg/100 g EP [31]. The high cholesterol concentrations in *Kabab Rubyan* (shrimp kebab; 359.17 mg/100 g) and *Marag Rubyan* (shrimp broth; 212.58 mg/100 g) are attributed to shrimp being the major ingredient in both food recipes. On a fresh weight basis, shrimp contain a high cholesterol concentration of 161 mg/100 g [32]. Cheese dishes (97.13–169.16 mg/100 g), cheeseburger based on beef (123.42 mg/100 g), *Shorbat Dajaj* based on chicken (103.47 mg/100 g), and *Samak Maleh* based on fish (86.75 mg/100 g) contained relatively high cholesterol levels. Among desserts, five dishes contained relatively low levels of cholesterol, *Baklawa* (4.42 mg/ 100 g), *Gherayba* (10.10 mg/ 100 g), *Qatayef* (dumpling filled with mixed nuts; 15.32 mg/ 100 g), *Halwa* (sweet wheat starch; 33.21 mg/ 100g) with the highest concentration of 56.60 mg/100 g in *Knafah*, that could be attributed to cheese and butter in the recipe. Other foods containing lower cholesterol levels were *Keema, Arayes, Madhrooba* (crushed wheat with lamb), and *Aish Fagae ma laham* (rice with truffles and lamb), all of which contained varying amounts of lamb. The cholesterol levels in meat-based foods analyzed in this study were higher than those reported for similar Saudi Arabian and Omani meat-based foods [25, 33].

### Fatty acid quality

Foods with low PUFA to SFA (P/S) ratios have been considered as undesirable in the diet, as they induce elevated levels of blood cholesterol, which contribute to an increased risk of CVD and cancers. In order to maintain a healthy cardiovascular status, P/S values higher than 0.45 are desirable [13, 34]. As presented in Table 3, the P/S ratios for meat-based foods (0.10–0.42), cheese dishes (0.01–0.04), desserts (*Gherayba* 0.12, *Knafah* 0.08, *Baklawa* 0.25,*Darabeel* 0.21, and *Zalabia* 0.19), cheeseburger (0.40), soup (*Shorbat Harees* (soup of peeled wheat with lamb) 0.05, and *Shorbat Dajaj* (chicken soup) 0.31) were all <0.45, which is considered nutritionally unhealthy. The remaining foods had higher P/S ratios (>0.45) and can thus be said to have good fat content for human nutrition concerning the balance between PUFA and SFA.

**Table 3.**
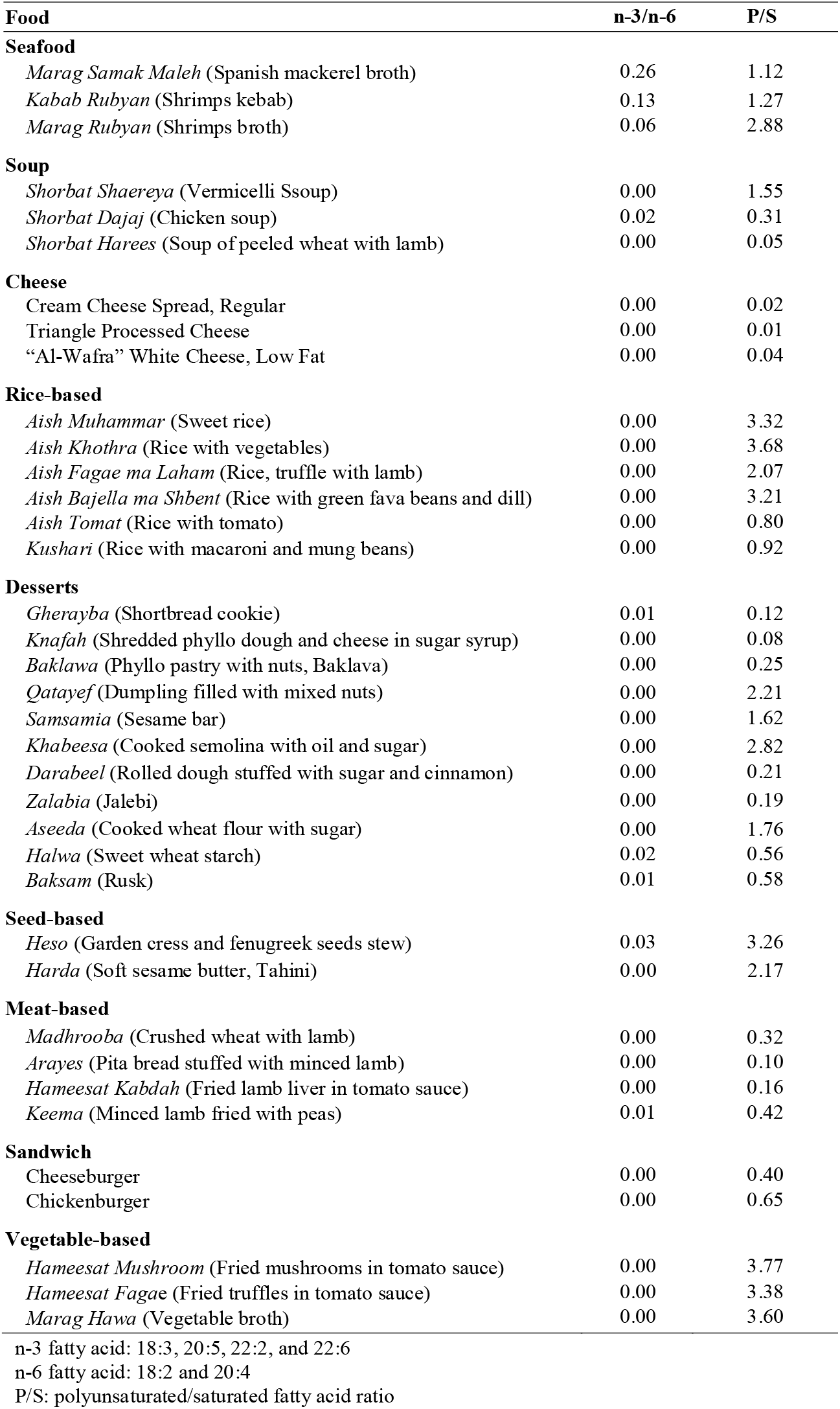
Fatty acid n-3/n-6 and polyunsaturated/saturated fatty acid ratio

| Food | n-3/n-6 | P/S |
| --- | --- | --- |
| **Seafood** |  |  |
| *Marag Samak Maleh* (Spanish mackerel broth) | 0.26 | 1.12 |
| *Kabab Rubyan* (Shrimps kebab) | 0.13 | 1.27 |
| *Marag Rubyan* (Shrimps broth) | 0.06 | 2.88 |
| **Soup** |  |  |
| *Shorbat Shaereya* (Vermicelli Soup) | 0.00 | 1.55 |
| *Shorbat Dajaj* (Chicken soup) | 0.02 | 0.31 |
| *Shorbat Harees* (Soup of peeled wheat with lamb) | 0.00 | 0.05 |
| **Cheese** |  |  |
| Cream Cheese Spread, Regular | 0.00 | 0.02 |
| Triangle Processed Cheese | 0.00 | 0.01 |
| “Al-Wafra” White Cheese, Low Fat | 0.00 | 0.04 |
| **Rice-based** |  |  |
| *Aish Muhammar* (Sweet rice) | 0.00 | 3.32 |
| *Aish Khothra* (Rice with vegetables) | 0.00 | 3.68 |
| *Aish Fagae ma Laham* (Rice, truffle with lamb) | 0.00 | 2.07 |
| *Aish Bajella ma Shbent* (Rice with green fava beans and dill) | 0.00 | 3.21 |
| *Aish Tomat* (Rice with tomato) | 0.00 | 0.80 |
| *Kushari* (Rice with macaroni and mung beans) | 0.00 | 0.92 |
| **Desserts** |  |  |
| *Gherayba* (Shortbread cookie) | 0.01 | 0.12 |
| *Knafah* (Shredded phyllo dough and cheese in sugar syrup) | 0.00 | 0.08 |
| *Baklava* (Phyllo pastry with nuts, Baklava) | 0.00 | 0.25 |
| *Qatayef* (Dumpling filled with mixed nuts) | 0.00 | 2.21 |
| *Samsamia* (Sesame bar) | 0.00 | 1.62 |
| *Khabeesa* (Cooked semolina with oil and sugar) | 0.00 | 2.82 |
| *Darabeel* (Rolled dough stuffed with sugar and cinnamon) | 0.00 | 0.21 |
| *Zalabia* (Jalebi) | 0.00 | 0.19 |
| *Aseeda* (Cooked wheat flour with sugar) | 0.00 | 1.76 |
| *Halwa* (Sweet wheat starch) | 0.02 | 0.56 |
| *Baksam* (Rusk) | 0.01 | 0.58 |
| **Seed-based** |  |  |
| *Heso* (Garden cress and fenugreek seeds stew) | 0.03 | 3.26 |
| *Harda* (Soft sesame butter, Tahini) | 0.00 | 2.17 |
| **Meat-based** |  |  |
| *Madhrooba* (Crushed wheat with lamb) | 0.00 | 0.32 |
| *Arayes* (Pita bread stuffed with minced lamb) | 0.00 | 0.10 |
| *Hameesat Kabdah* (Fried lamb liver in tomato sauce) | 0.00 | 0.16 |
| *Keema* (Minced lamb fried with peas) | 0.01 | 0.42 |
| **Sandwich** |  |  |
| Cheeseburger | 0.00 | 0.40 |
| Chickenburger | 0.00 | 0.65 |
| **Vegetable-based** |  |  |
| *Hameesat Mushroom* (Fried mushrooms in tomato sauce) | 0.00 | 3.77 |
| *Hameesat Fagae* (Fried truffles in tomato sauce) | 0.00 | 3.38 |
| *Marag Hawa* (Vegetable broth) | 0.00 | 3.60 |

n-3 fatty acid: 18:3, 20:5, 22:2, and 22:6
n-6 fatty acid: 18:2 and 20:4

A balanced n-3/n-6 fatty acid ratio has positive effects on decreasing the risk of CVD and cancers [35]. The n-3/n-6 fatty acid ratios below 4.0 were recommended by the Department of Health of the United Kingdom [36]. The n-3/n-6 ratio of all foods examined in this study (0.00 and 0.26) were within the recommendations (Table 3). Although, recent studies by Marventano et al. and FAO/WHO [23, 37] have suggested that there might not be any impact of n-3/n-6 fatty acid ratio on human health.

An atherogenic and/or thrombogenic food is one that has the potential to induce platelet aggregation, which might stimulate CVD, and is thus considered less nutritionally beneficial. The SFAs, C12:0, C14:0, and C16:0 are considered atherogenic and thrombogenic as many studies have shown that they have a cholesterol-raising effect that may induce platelets aggregation [38]. Thus, foods with higher concentrations of these fatty acids have higher atherogenic (AI) and thrombogenic indices (TI).

AI and TI values were calculated in the present study, these values indicate the potency of foods to stimulate platelet aggregation, and lower values indicate better nutritional quality of food. AI and TI values were the highest for cheese dishes, and ranged from 2.11 to 2.84, and 3.18 to 4.03; respectively (Table 4). The AI and TI values were variable for desserts, AI values ranged from 0.17 in *Khabeesa* and 2.31 in *Knafah*, while TI values ranged from 0.37 in *Khabeesa* and 3.29 in *Knafah*. The high AI and TI values for *Knafah* were attributed to the presence of around 50% C14:0 and C16:0 SFA. Soup was the third highest food group with AI ranging from 0.36–1.12, and TI from 0.66–2.61. AI ranged between 0.61–0.78 and TI between 1.30–1.58 for sandwiches, 0.51–0.66 and 1.29–2.07 for meat-based food, 0.18–0.55 and 0.32–0.58 for seafood, and 0.14–0.70 and 0.33–1.13 for rice-based food. A low AI of 0.15 was recorded for all vegetable-based foods, as well as low TI values varied between 0.33 and 0.35. AI for seed-based *Harda* and *Heso* (garden cress and fenugreek seeds stew) was the lowest among all foods investigated in this study, with values of 0.12 and 0.14, respectively; and the lowest TI among all foods was 0.29 for *Heso*. This is attributed to low C14:0 and C16:0 SFA and high MUFA and PUFA levels in *Harda* and *Heso*.

**Table 4.**
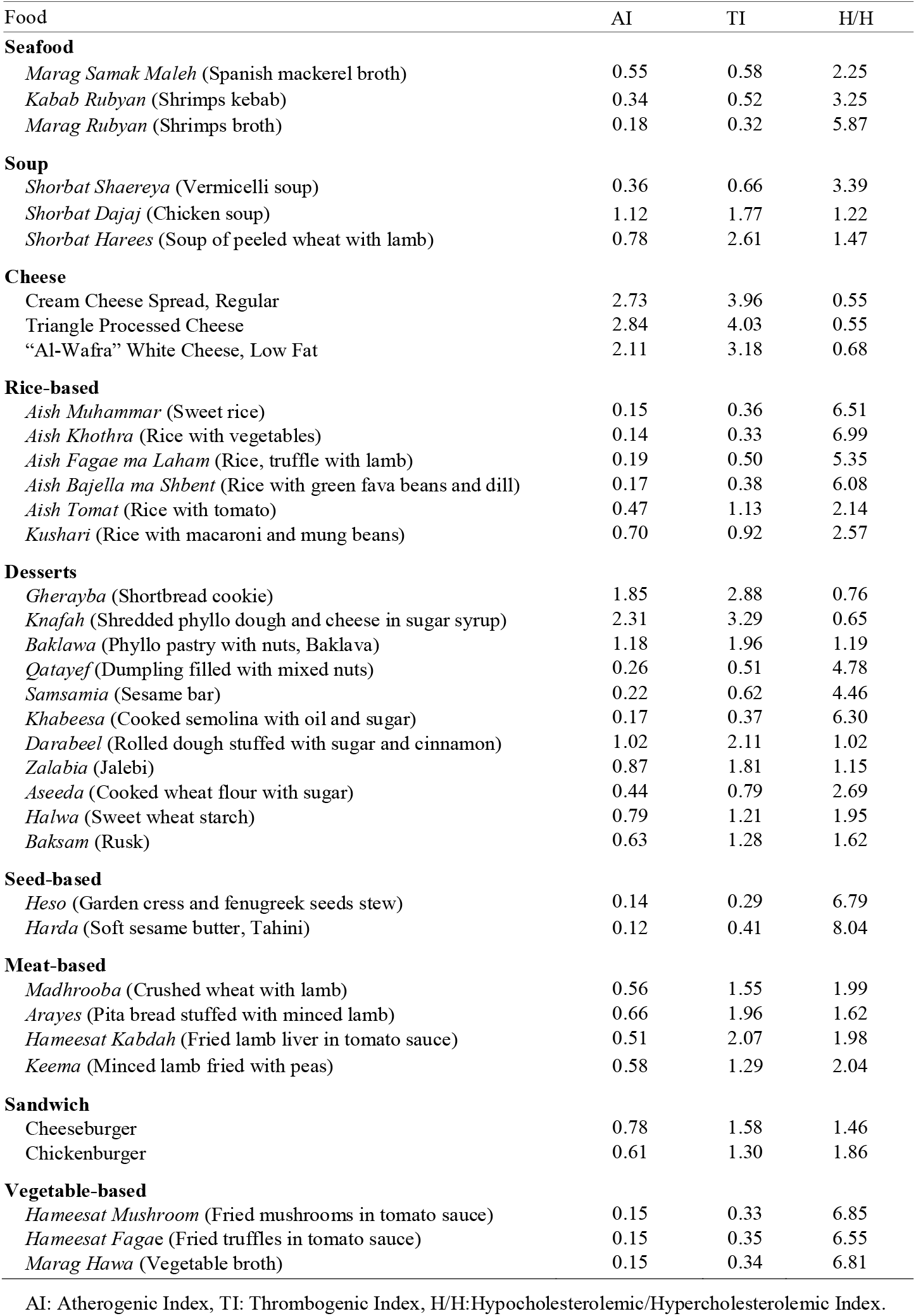
Fatty acid quality of 37 local foods of the State of Kuwait using atherogenic, thrombogenic and hypocholesterolemic/hypercholesterolemic indices.

| Food | AI | TI | H/H |
| --- | --- | --- | --- |
| **Seafood** |  |  |  |
| *Marag Samak Maleh* (Spanish mackerel broth) | 0.55 | 0.58 | 2.25 |
| *Kabab Rubyan* (Shrimps kebab) | 0.34 | 0.52 | 3.25 |
| *Marag Rubyan* (Shrimps broth) | 0.18 | 0.32 | 5.87 |
| **Soup** |  |  |  |
| *Shorbat Shaereya* (Vermicelli soup) | 0.36 | 0.66 | 3.39 |
| *Shorbat Dajaj* (Chicken soup) | 1.12 | 1.77 | 1.22 |
| *Shorbat Harees* (Soup of peeled wheat with lamb) | 0.78 | 2.61 | 1.47 |
| **Cheese** |  |  |  |
| Cream Cheese Spread, Regular | 2.73 | 3.96 | 0.55 |
| Triangle Processed Cheese | 2.84 | 4.03 | 0.55 |
| “Al-Wafra” White Cheese, Low Fat | 2.11 | 3.18 | 0.68 |
| **Rice-based** |  |  |  |
| *Aish Muhammad* (Sweet rice) | 0.15 | 0.36 | 6.51 |
| *Aish Khothra* (Rice with vegetables) | 0.14 | 0.33 | 6.99 |
| *Aish Fagae ma Laham* (Rice, truffle with lamb) | 0.19 | 0.50 | 5.35 |
| *Aish Bajella ma Shbent* (Rice with green fava beans and dill) | 0.17 | 0.38 | 6.08 |
| *Aish Tomat* (Rice with tomato) | 0.47 | 1.13 | 2.14 |
| *Kushari* (Rice with macaroni and mung beans) | 0.70 | 0.92 | 2.57 |
| **Desserts** |  |  |  |
| *Gherayba* (Shortbread cookie) | 1.85 | 2.88 | 0.76 |
| *Knafah* (Shredded phyllo dough and cheese in sugar syrup) | 2.31 | 3.29 | 0.65 |
| *Baklava* (Phyllo pastry with nuts, Baklava) | 1.18 | 1.96 | 1.19 |
| *Qatayef* (Dumpling filled with mixed nuts) | 0.26 | 0.51 | 4.78 |
| *Samsamia* (Sesame bar) | 0.22 | 0.62 | 4.46 |
| *Khabeesa* (Cooked semolina with oil and sugar) | 0.17 | 0.37 | 6.30 |
| *Darabeel* (Rolled dough stuffed with sugar and cinnamon) | 1.02 | 2.11 | 1.02 |
| *Zalabia* (Jalebi) | 0.87 | 1.81 | 1.15 |
| *Aseeda* (Cooked wheat flour with sugar) | 0.44 | 0.79 | 2.69 |
| *Halwa* (Sweet wheat starch) | 0.79 | 1.21 | 1.95 |
| *Baksam* (Rusk) | 0.63 | 1.28 | 1.62 |
| **Seed-based** |  |  |  |
| *Heso* (Garden cress and fenugreek seeds stew) | 0.14 | 0.29 | 6.79 |
| *Harda* (Soft sesame butter, Tahini) | 0.12 | 0.41 | 8.04 |
| **Meat-based** |  |  |  |
| *Madhrooba* (Crushed wheat with lamb) | 0.56 | 1.55 | 1.99 |
| *Arayes* (Pita bread stuffed with minced lamb) | 0.66 | 1.96 | 1.62 |
| *Hameesat Kabdah* (Fried lamb liver in tomato sauce) | 0.51 | 2.07 | 1.98 |
| *Keema* (Minced lamb fried with peas) | 0.58 | 1.29 | 2.04 |
| **Sandwich** |  |  |  |
| Cheeseburger | 0.78 | 1.58 | 1.46 |
| Chickenburger | 0.61 | 1.30 | 1.86 |
| **Vegetable-based** |  |  |  |
| *Hameesat Mushroom* (Fried mushrooms in tomato sauce) | 0.15 | 0.33 | 6.85 |
| *Hameesat Fagae* (Fried truffles in tomato sauce) | 0.15 | 0.35 | 6.55 |
| *Marag Hawa* (Vegetable broth) | 0.15 | 0.34 | 6.81 |

AI: Atherogenic Index, TI: Thrombogenic Index, H/H: Hypocholesterolemic/Hypercholesterolemic Index.

Contrary to AI and TI, foods with higher H/H indices are more nutritionally desirable [39]. The highest H/H (8.04) was recorded for *Harda*. The H/H range for the different foods examined in this study from the highest to lowest values were: rice-based foods (2.14–6.99), vegetable-based foods (6.55–6.85), *Heso* (6.79), seafood (2.25–5.87), desserts (0.65–4.78, except *Khabeesa* with H/H 6.29), soup (1.22–3.39), meat-based foods (1.62–2.04), sandwiches (1.46–1.86), and cheese dishes with the lowest H/H values (0.55–0.68). The AI, TI, and H/H values may indicate that the consumption of seed-based, vegetable-based, rice-based, and seafood dishes analyzed in the present study is potentially healthier, in terms of the fatty acid quality, for humans than that of cheese, desserts, sandwiches, and meat-based foods.

## Conclusions

The results of this study revealed a high percentage of unsaturated fatty acids, predominantly oleic and linoleic acids, and favorable indices such as P/S, AI, TI, and H/H for seafood dishes, rice-based, seed-based, and vegetable-based foods, suggesting that these foods can be consumed as part of a healthy diet. The cholesterol content was high in two shrimp-based seafood dishes; hence, it is preferable that these foods be consumed with caution by people with high blood cholesterol levels. The TFA content was low in all foods examined, at levels that do not represent any known significant health risk. Interestingly, all foods showed n-3/n-6 fatty acid ratios within the nutritional recommendations. The highest SFA content (predominantly C16:0 and C14:0) was recorded in cheese and some ready-made desserts including *Knafah, Gherayba, Baklawa*, and *Zalabia*. It is worth mentioning that these desserts are also popular among the Middle East and Mediterranean populations. The same desserts showed low fatty acid quality based on their P/S, AI, TI, and H/H values; thus, they should be consumed in moderation or other strategies should be adopted, such as modifying the composition of these foods, by for example substituting ghee or butter with vegetable oil. Published data on foods with similar structure could not be compared with our data, since other studies reported fatty acid data on a dry weight basis. The fatty acid and cholesterol data obtained in the present study can be compiled to national food composition tables and incorporated in international databases such the United States Department of Agriculture (USDA) Nutrient Database for Standard Reference and EuroFIR FoodEXplorer to fill in missing data on fatty acids and cholesterol composition of local foods. Furthermore, data of the present study could be used in epidemiological and nutrition-health related studies aim at assessing population fatty acid nutritional status, and hence studying the associations between dietary fatty acids and the etiology of CVD and/or other nutrition-related diseases in humans.

## Data Availability

All data referred to in the manuscript are submitted with the manuscript

## Supplementary material

S1 Table. Detailed ingredients for 37 local Kuwaiti foods.

S2 Table. Fatty acid composition (g/100 g EP) for 37 local Kuwaiti foods.

## Acknowledgments

The authors gratefully acknowledge the Ministry of Health of the State of Kuwait (MOH) for their financial support, and the Management of Kuwait Institute for Scientific Research (KISR) for their support and encouragement.

## Author contributions

**Hanan A. Al-Amiri:** Conceptualization, Validation, Data Analysis, Visualization, Supervision, Project Administration, Funding acquisition, Writing-Original Draft, Writing-Reviewing & Editing

**Nisar Ahmed, Tahani Al-Sharrah**: Methodology, Investigation, Resources.

