## Supplemental Table 1 for "Fatty acid profiles, cholesterol composition, and nutritional quality indices of 37 commonly consumed local foods in Kuwait in relation to cardiovascular health"

| **S1 Table. Detailed ingredients for 37 local Kuwaiti foods** |  |
| --- | --- |
| ***Local Name* (Common Name)** | **Ingredients (%)** |
| **Seafood** |  |
| *Marag Samak Maleh* (Spanish Mackerel Broth) | Spanish Mackerel Fish (33), Tomato (10), Onion (9), Tomato |
|  | Paste(5), Coriander (5), Dill (5), Corn Oil (3), Garlic (2), Dried |
|  | Lemon (1), Mixed Spices^a^ (1), Water (28) |
| *Kabab Rubyan* (Shrimps Kebab) | Shrimps (60), Rice Flour (15), Onion (14), Coriander (8), Corn |
|  | Oil (1), Mixed Spices (1), Salt (0.4), Garlic (0.3), Turmeric (0.2) |
|  | Dried Lemon (0.2), Cardamom (0.2), Clove (0.2) |
| *Marag Rubyan* (Shrimps Broth) | Shrimp (36), Tomato (13), Onion (8), Tomato Paste (5), Salt (1) |
|  | Mixed Spices (1), Corn Oil (0.4), Coriander (0.3), Water (36) |
| **Soup** |  |
| *Shorbat Shaereya* (Vermicelli Soup) | Vermicelli (12), Carrots (8), Tomato (7), Salt (2), Parsley (0.3) |
|  | Water (70) |
| *Shorbat Dajaj* (Chicken Soup) | Chicken breast (48), Milk (14),Wheat Flour (2), Butter (2), Salt |
|  | (1), Starch (1), Black Pepper (0.2), Water (32) |
| *Shorbat Harees* (Soup of Peeled Wheat with Lamb) | Peeled Wheat (15), Lamb (14), Tomato Paste (8), Water (62), |
|  | Salt (1) |
| **Cheese** |  |
| Cream Cheese Spread, Regular* |  |
| Triangle Processed Cheese* |  |
| “Al-Wafra” White Cheese, Low Fat* |  |
| **Rice-based** |  |
| *Aish Muhammar* (Sweet Rice) | Rice (23), Sugar (20), Corn Oil (2), Rosewater (1), Saffron |
|  | (0.1), Water (53) |
| *Aish Khothra* (Rice with Vegetables) | Rice (21), Eggplant (17), Potato (14), Marrow (12), Onion (8), |
|  | Tomato (6), Salt (0.4), Spices (0.3), Corn Oil (0.2), Water (22) |
| *Aish Fagae ma Laham* (Rice, Truffle with Lamb) | Rice (16), Truffle (11), Lamb (9), Onion (9), Salt (1), Corn Oil |
|  | (0.4), Black Pepper (0.2),Water (53) |
| *Aish Bajella ma Shbent* (Rice with Green | Rice (40), Green Fava Beans (21), Dill (7), Onion (6), Salt (1), |
| Fava Beans and Dill) | Corn Oil (0.3), Water (25) |
| *Aish Tomat* (Rice with Tomato) | Rice (39), Tomato (6),Onion (4), Mixed Spices (0.1), Corn Oil |
|  | (0.4), Water (52) |
| *Kushari* (Rice with Macaroni and Mung Beans) | Rice (21), Macaroni Pasta (16), Mung Beans (9),Vermicelli (7), |
|  | Onion (6), Corn Oil (4), Salt (1), Mixed Spices (0.4), Water (35) |
| **Desserts** |  |
| *Gherayba* (Shortbread Cookie)*** | Brown Wheat Flour, Corn Flour, Sugar, Ghee, Cardamom |
| *Knafah* (Shredded Phyllo Dough and Cheese in | Phyllo Dough, Butter, Mozzarella Cheese,Sugar Syrup (Sugar, |
| Sugar Syrup)* | Rosewater) |
| *Baklawa* (Phyllo Pastry with Nuts, Baklava)* | Phyllo Pastry, Egg White, Butter, Pistachio Nuts, Walnut, |
|  | Caster Sugar, Sugar Syrup |
| *Qatayef* (Dumpling Filled with Mixed Nuts^b^)* | Semolina, Starch Corn, Ghee (for frying), Nuts, Sugar Syrup |
| *Samsamia* (Sesame Bar)* | Sesame Kernel Seeds, Molasses, Sugar |
| **S1 Table. (Continued)** |  |
| ***Local Name* (Common Name)** | **Ingredients (%)** |
| *Khabeesa* (Cooked Semolina with Oil and Sugar) | Semolina (27), Sugar (19), Corn Oil (6), Cardamom (0.2), |
|  | Water (48) |
| *Darabeel* (Rolled Dough Stuffed with Sugar and | Brown Wheat Flour, White Wheat Flour, Ghee, Sugar, |
| Cinnamon* | Rosewater, Cardamom, Cinnamon |
| *Zalabia* (Jalebi)* | White Wheat Flour, Sugar, Ghee, Saffron, Cardamom, Yeast, |
|  | Water, Sugar Syrup |
| *Aseeda* (Cooked Wheat Flour with Sugar) | Sugar (28), Wheat Flour (15), Corn Oil (5), Water (53) |
| *Halwa* (Sweet Wheat Starch)* | Wheat Starch, White Sugar, Brown Sugar, Ghee, Nuts |
|  | Rosewater, Saffron |
| *Baksam* (Rusk)* | Brown Wheat Flour, Ghee, Sugar, Yeast, Cardamom |
| **Seed-based** |  |
| *Heso* (Garden Cress and Fenugreek Seeds Stew) | Garden Cress Seeds (9), Fenugreek Seeds (5), Spices (Ginger, |
|  | Cinnamon, Cardamom, Black Pepper, Turmeric) (4), Sugar (4) |
|  | Corn Oil (4), Water (74) |
| *Harda* (Soft Sesame Butter, Tahini)* | Sesame Kernel Seeds |
| **Meat-based** |  |
| *Madhrooba* (Crushed Wheat with Lamb) | Crushed Wheat (17), Lamb (17), Onion (13), Tomato (11), |
|  | Tomato Paste (5), Salt (1), Mixed Spices (0.4), Dried Lemon |
|  | (0.3), Black Pepper (0.2), Water (35) |
| *Arayes* (Pita Bread Stuffed with Minced Lamb)* | Pita Bread, Minced Lamb , Tomato,Tomato Paste, Onion, Oil |
|  | Parsley, Salt, Mixed Spices, Garlic |
| *Hameesat Kabdah* (Fried Lamb Liver in Tomato | Lamb Liver (90), Onion (6), Tomato Paste (2), Corn Oil (1), |
| Sauce) | Mixed Spices (0.4), Salt (0.3) |
| *Keema* (Minced Lamb with Peas) | Minced Lamb (61), Peas (16),Onion (14),Tomato Paste |
|  | (8), Corn Oil (1), Mixed Spices (0.3), Salt (0.3) |
| **Sandwich** |  |
| Cheeseburger* | Beef Meat, Bun Bread, Sliced Cheese, Tomato, Lettuce, Onion |
|  | Pickles, Ketchup, Mayonnaise |
| Chicken burger* | Chicken "Fillet", Bun Bread, Sliced Cheese, Lettuce, Onion, |
|  | Pickles, Ketchup, Mayonnaise |
| **Vegetable-based** |  |
| *Hameesat Mushroom* (Fried Mushroom in | Mushroom (35), Onion (32), Tomato (27), Tomato Paste (3), |
| Tomato Sauce) | Corn Oil (2), Salt (2), Mixed Spices(1), Garlic (0.4) |
| *Hameesat Faga*e (Fried Truffle in Tomato Sauce) | Truffle (41), Tomato (25), Tomato Paste (21), Onion (9), Corn |
|  | (9), Corn Oil (3), Salt (1), Mixed Spices (1) |
| *Maraq Hawa* (Vegetable Broth) | Eggplant (19), Potato (15), Green Beans (12), Onion (10), |
|  | Tomato (10), Zucchini (9), Butternut Squash (6), Tomato Paste |
|  | (3), Corn Oil (0.5), Mixed Spices (0.2), Water (16) |
| *Ready-made purchased foods |  |

^a^Mixed Spices: black pepper, cayenne pepper, coriander, cardamom, cloves, cumin, cinnamon, all ground and in equal amounts.

^b^Mixed Nuts: hazelnut and walnut with raisins
